# Genetic evidence for protective effects of smoking and drinking behavior on Parkinson’s disease: A Mendelian Randomization study

**DOI:** 10.1101/2020.04.20.20073247

**Authors:** Carmen Domínguez-Baleón, Jue-Sheng Ong, Clemens R. Scherzer, Miguel E. Rentería, Xianjun Dong

## Abstract

**Background:** Observational studies have identified correlations between environmental and lifestyle factors and Parkinson’s disease (PD). However, the causal direction of many of these relationships remains unclear.

**Objective:** To infer causal relationships between smoking and alcohol intake and PD.

**Methods:** We use a two-sample Mendelian randomization (MR) experimental design to infer causal relationships between smoking (*initiation, age of initiation, heaviness*, and *cessation*) and alcohol (*drinks per week*) consumption as exposure variables and PD as the health outcome. We also conduct sensitivity analyses, including testing for pleiotropic effects MR-Egger and MR-PRESSO, and *multivariable MR* to jointly model the effects of drinking and smoking behavior on PD risk.

**Results:** Both *alcohol intake* (OR = 0.69; 95% CI 0.56-0.86; p=0.001). and *smoking cessation* (comparing *current* vs. *former smokers*) (IVW OR = 0.39; 95% CI 0.22 to 0.69; p=0.001) were causally associated with a reduced risk of PD. In addition, our multivariable MR results provide additional assurance that the causal association between *drinks per week* and PD is unlikely due to confounding by smoking behavior.

**Conclusion:** Our findings support the role of smoking as a protective factor against PD, but only when comparing *current vs. former smokers*. Increased alcohol intake also had a protective effect over PD risk, and the *alcohol dehydrogenase 1B* (*ADH1B*) locus is a candidate for further investigating the mechanisms underlying this association.

## INTRODUCTION

Parkinson’s disease (PD) is a complex genetic disease, and growing evidence exists that lifestyle and environmental factors ^1^ and multiple genetic variants modulate both its susceptibility ^2,3^ and progression ^4,5^. Observational studies have uncovered links between Parkinson’s disease (PD) risk and environmental factors. For instance, exposure to pesticides ^6,7^ and a history of melanoma^8^ have been associated with increased risk of PD, whereas alcohol intake ^9,10,11^, smoking ^12,13,14^, caffeine consumption ^15,16^, and educational attainment^17^ are correlated with reduced risk.^1,18^ However, causality cannot be inferred purely from association studies, as these relationships may be due to factors such as confounding variables or reverse causation.^19,20^ A traditional approach to infer causality is the Randomized Controlled Trial (RCT), where participants are randomly allocated to either a control or a treatment group, thus reducing bias.^21^ The major downside of an RCT, however, is the ethical and health concerns of subjecting participants to exposures that might be detrimental to their health, such as smoking, drinking, or pesticides.^22,23,24^

Mendelian randomization (MR) is a promising alternative methodology that overcomes these difficulties by using genetic variants (e.g., single nucleotide polymorphisms, or SNPs) as instruments to infer causality between environmental exposures and health outcomes in a way that is independent of confounding factors. This method takes advantage of the fact that genetic variants are randomly distributed in the population just as participants would randomly be allocated in an RCT.^20,22,24,25^ A *two-sample MR* experimental design requires summary statistics from two independent genome-wide association studies (GWAS), one for the exposure and one for the outcome variables. It examines whether the SNPs (*S*) that influence the exposure (*E*) also affect the outcome (*O*), but not the other way around. This way, MR facilitates the inference of a direct causal relationship between the exposure and the outcome (See **Figure 1**).^22,23^

**Figure 1.**
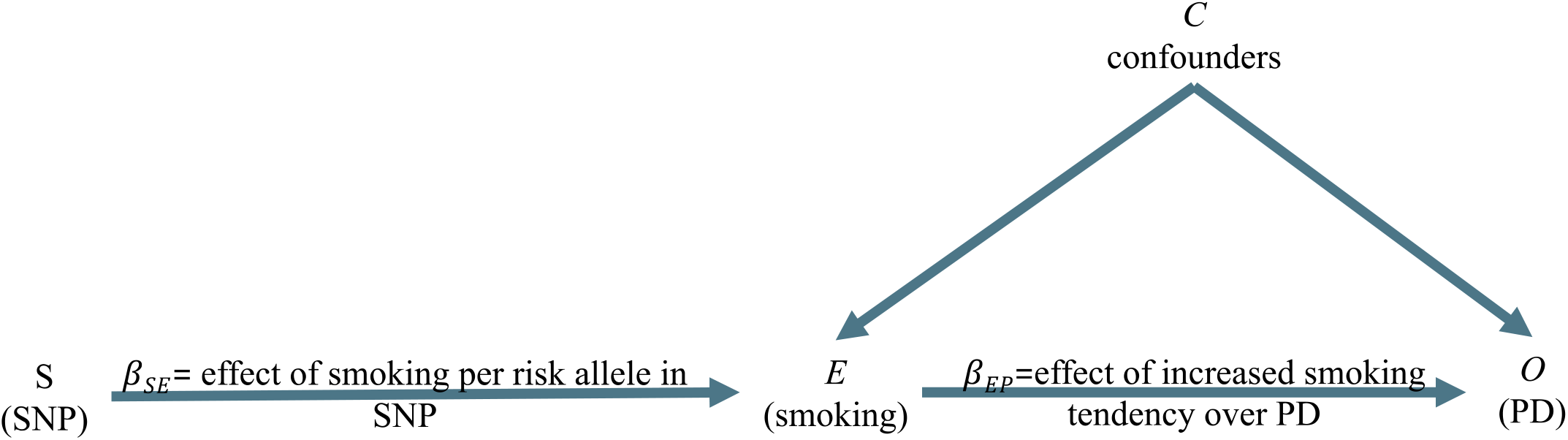
Directed acyclic graph of the effect of smoking on PD using genetic variants (SNPs) as proxies. (Adapted from Lawlor et al., 2008^22^). A SNP (S) is associated with the exposure, for example, smoking (E), and therefore the SNP (S) can be used as an instrument to determine the effect of smoking (*E*) on PD, which is the outcome of interest (*O*). An association between the smoking-risk-increasing SNP (*S*) and elevated risk of PD is consistent with a causal relationship between *E* and *O*, if three assumptions are met: (1) The SNP (*S*) must be strongly associated with smoking (*E*); (2) The SNP (*S*) must be independent of any confounding factors (*C*) that may be influencing PD (*O*) and (3) The SNP (S)’s association with PD (*O*) is explained only via smoking (*E*). SNP, single nucleotide polymorphism.

The inverse association of PD with smoking and alcohol intake has been consistently reported in the literature.^1,9,10,11,12,13,14,18^ A recent meta-analysis across 61 case-control and eight cohort studies by Li *et al*.^13^ estimated a 0.59 (95% CI 0.56-0.62) pooled relative risk (RR) of PD in smokers. Furthermore, another meta-analysis of 32 studies by Zhang *et al*.^10^ estimated a RR of 0.78 (95% CI 0.67-0.92) when comparing participants with the highest versus the lowest levels of alcohol intake while also controlling for the effects of smoking and caffeine intake. Notably, between-study heterogeneity has been observed, which implies that studies in larger samples might be needed to confirm the direction of the causality of this association.

Recently, Mendelian randomization has been used to try to infer causality between PD and smoking and drinking. Nalls *et al*.^27^ reported little evidence for a causal relationship between current tobacco use and PD risk, despite finding a significant inverse genetic relationship between the two traits, estimated using LD score regression. Interestingly, Noyce *et al*.^28^ conducted an extensive study with 401 exposure traits, including smoking and drinking. Despite reporting negative causal associations, the authors did not explore in further detail with sensitivity analyses to evaluate biases due to horizontal pleiotropy (i.e., a situation where candidate SNP instruments might be associated with the outcome through pathways independent of the exposure of interest). Hence it was unclear whether key MR assumptions were robustly satisfied when interpreting those findings.

Recent studies by the GWAS & Sequencing Consortium of Alcohol and Nicotine use (GSCAN)^26^ and the International Parkinson’s Disease Genomics Consortium^27^ (IPDGC) have identified or confirmed hundreds of genetic variants robustly associated with smoking behavior,^26^ alcohol intake^26^ and Parkinson’s disease susceptibility.^27^ The GSCAN study involved up to 1.2 million participants, while the IPDGC meta-analysis comprised 35,24037,688 PD cases, 18,618 proxy-cases (defined as first-degree relatives of a PD patient), and 900,2381.4 million controls. The present study leverages the availability of GWAS summary statistics from these new studies to expand on previous MR investigations. In particular, we examine the causal effects of five lifestyle variables (*number of alcoholic drinks per week, smoking initiation, heaviness, cessation*, and *age of initiation*) on PD risk. Notably, given that smoking and alcohol intake are both phenotypically and genetically correlated, we also conduct a multivariable MR analysis to control for horizontal pleiotropy on substance use behavior, thus providing a better adjusted and more robust inference model.

## RESULTS

We observed statistically-significant inverse (i.e., protective) effects for the *number of drinks per week* and *smoking cessation* (which compares *current vs. former smokers*) on PD risk. Besides, despite displaying consistent effect estimates across all MR methods used, *smoking initiation* (*ever vs. never being a regular smoker*) did not reach statistical significance. The results for these three phenotypes are summarized in **Figure 2** and described below.

**Figure 2.**
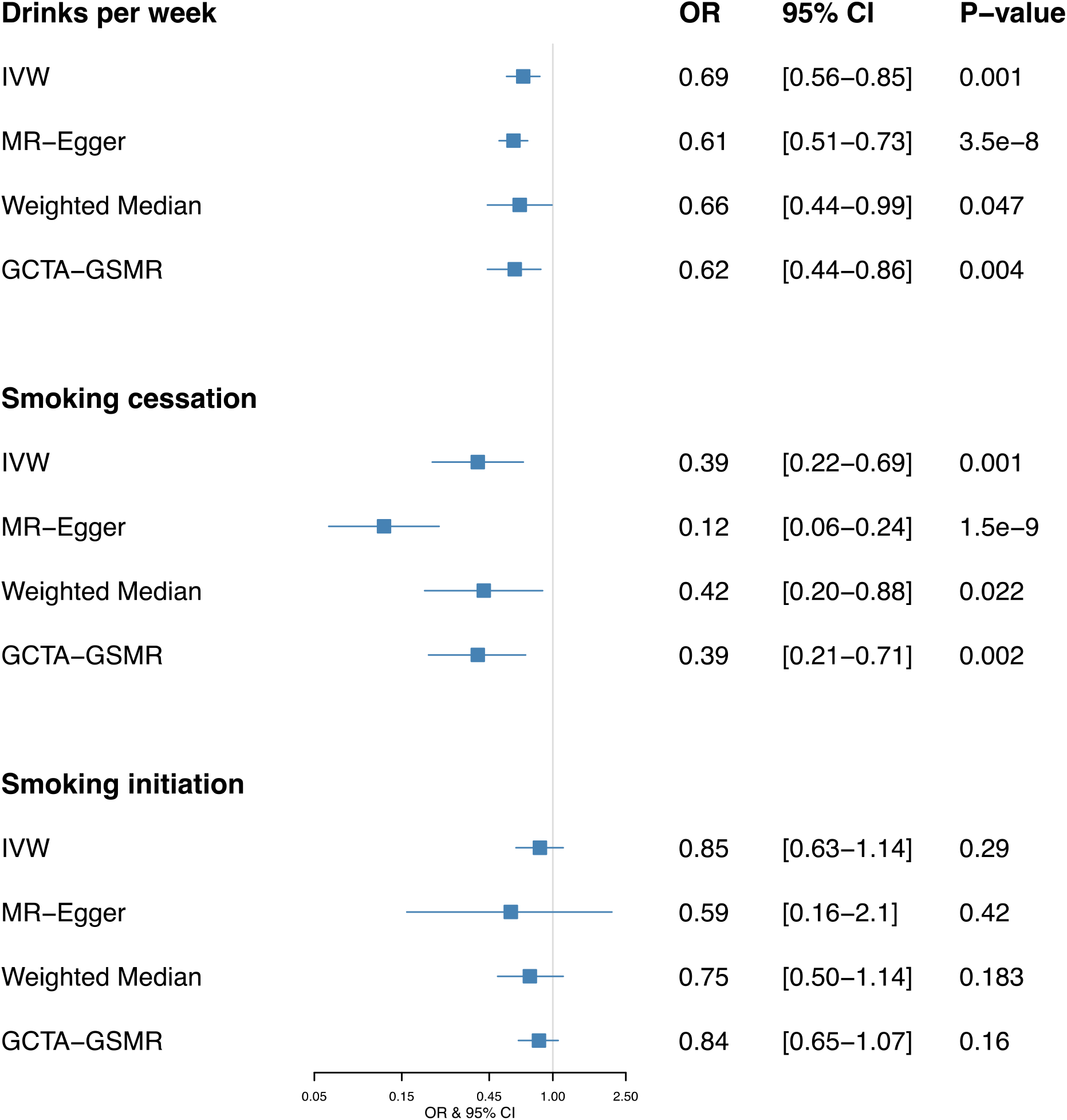
Forest plot showing OR and 95% confidence intervals of MR effect estimates. OR values for binary traits (*smoking cessation* and *smoking initiation)* are expressed in *per doubling of odds* units.

### Alcohol consumption

Horizontal pleiotropy analysis with MR-PRESSO identified two potentially pleiotropic outliers (rs1260326 and rs113589236) which which have previously been associated with other traits, such as triglyceride levels^29^ and epithelial ovarian cancer.^30^ After excluding outliers, our MR estimates showed that the genetic tendency to have more *drinks per week* is causally associated with a reduced PD risk. The OR from the IVW estimate was 0.69 (95% CI 0.56-0.85; p=0.001) (See **Figure 2**), and the effect estimate from the MR-Egger regression was 0.61 (95% CI 0.51-0.73; p=3.5e-8) with a negligibly small intercept of 0.005 (p=0.259) indicating no presence of horizontal pleiotropy. Estimates derived from GCTA-GSMR yielded very similar findings: OR was 0.62 (95% CI 0.44-0.86; p=0.004). There was no evidence of heterogeneity of effect estimates between variants (Q statistic = 36.96, I^2^=95.9%, p=0.52). We did not find evidence for unbalanced pleiotropy via manual inspection in the MR funnel plots (See **Supplementary Materials**). Notably, the rs1229984 variant on the *alcohol dehydrogenase 1B* (*ADH1B*) locus had the greatest association with *drinks per week*. It is known that individuals with one or two A alleles (i.e., AG or AA genotypes) at rs1229984 are more likely to find drinking unpleasant and thus have a reduced risk for alcoholism.^31,32^ After removing this SNP and re-running all analyses, the resulting estimate was only slightly attenuated towards the null and the point estimate remained in the same direction [0.74 (95% CI 0.46-1.19; p=0.219)] - suggesting that our results are unlikely to be solely driven by the rs1229984 variant, and that alcohol intake confers a robust protective effect over PD risk. However, the mechanism by which greater alcohol intake reduces PD risk requires further investigation.

We observed no evidence of reverse causality in our bidirectional MR analysis. Parkinson’s disease predisposition did not influence *drinks per week*, with an IVW estimate of 1.005 (95% CI 0.998 - 1.01; p=0.122), and a GSMR estimate of 1.027 (95% CI 0.993 - 1.062; p=0.117) per doubling of odds on PD. A *doubling of odds* refers to a doubling in the prevalence of the binary exposure and was first introduced by Gage *et al*.^33^ to facilitate the interpretation of effect estimates. This result indicates that the effect of *drinks per week* on PD is not bidirectional, as PD does not have a significant effect over *drinks per week*. Finally, multivariable MR analysis found no attenuation of the effect of alcohol on PD risk [i.e., MVMR OR 0.64 (95% CI 0.42 to 0.99) per standard deviation increase in the number of drinks per week] when the genetic effect of alcohol SNPs on smoking behavior, obesity and education attainment have been taken into consideration.

### Smoking cessation

MR-PRESSO identified a single SNP outlier (rs9607805, known to be associated with neuroticism^34^), which was excluded from the analysis. IVW results showed that current smokers have a lower risk of PD in contrast to former smokers. The OR from the IVW estimate was 0.39 (95% CI 0.22 to 0.69; p=0.001) per doubling of odds (See **Figure 2**), whereas the MR-Egger estimate was 0.12 (95% CI 0.06 - 0.24; p=1.5e-9) per doubling of odds with an intercept (in log(OR)) of 0.039, p=1.5e-5, which suggests that even after excluding the outlier, weak unbalanced horizontal pleiotropy is still present across all SNPs. Moderate evidence of heterogeneity among variants (Q statistics = 1.5, I^2^=75.8%, p=0.91) was observed. Results from the GSMR analysis were very similar to IVW, with OR=0.39 (95% CI 0.21 - 0.71; p=0.002) per doubling of odds. Given the existing discrepancies in results from IVW/GSMR and MR-Egger it is not possible to rule out the possibility that pleiotropic effects may influence the association between *being a current smoker* and PD. Bidirectionality analysis showed that the effect of predisposition to Parkinson’s disease over *smoking cessation* is very close to the null, with a non-significant p-value and an IVW estimate of 1.003 (95% CI 0.99 to 1.009; p=0.170) per doubling of odds. GSMR analysis yielded similar results: 1.026 (95% CI 0.98 - 1.07; p=0.244), per doubling of odds of PD on *being a current smoker*. Importantly, the marginal multivariable MR OR estimate could not be meaningfully interpreted due to the low number of SNP instruments, which yields an extensive confidence interval [OR 0.33 (95% CI 00 to >10)].

### Ever vs never being a regular smoker

Despite not reaching statistical significance, the predisposition to *ever being a regular smoker* (as opposed to never initiating smoking) was consistently associated with lower PD risk. The OR IVW estimate was 0.85 (95% CI 0.63 to 1.14; p=0.29) per doubling of odds and the GSMR estimate was 0.84 (95% CI 0.66 - 1.07; p=0.16) per doubling of odds (See **Figure 2**). Moreover, the MR-Egger estimate was 0.59 (95% CI 0.16 to 2.1; 0=0.42) per doubling of odds with minimal evidence of heterogeneity between variants (Q statistic = 119.62, I^2^=40%, p=0.38). Besides, the symmetrical distribution of variants on the funnel plot (see **Supplementary Materials**) suggests no horizontal pleiotropy. Bidirectional MR analyses found no evidence of reverse causality, with an IVW estimate of 1.0007 (95% CI 0.995 - 1.005, p=0.900) per doubling of odds and a GSMR estimate of 1.003 (95% CI 0.97 - 1.024, p=0.83) per doubling of odds. Similar to *smoking cessation*, the estimated OR confidence intervals from the multivariable MR analysis [2.11 (95% CI 0.39 to 11.51)] was too wide to make any reliable inference.

## DISCUSSION

In the present study, we provide genetic evidence to support a protective role of moderate alcohol intake against PD risk PD 0.69 (95% CI 0.56-0.86; p=0.001) Both the effect estimate and direction of causal association were consistent (OR 0.74 (95% CI 0.46-1.19; p=0.219) even after removing the rs1229984 variant, which had the strongest association with *drinks per week*. Therefore, we can conclude that a single SNP does not exclusively drive the observed causal association. Considering that this variant is both implicated in alcohol metabolism and strongly associated with alcohol intake, it is a promising candidate for investigating the mechanism by which alcohol drinking influences PD risk.

The ADH1B protein metabolizes ethanol into acetaldehyde during alcohol metabolism, and the risk allele rs1229984 (A) produces a substitution of Arginine for a Histidine that changes affinity for the NAD+ coenzyme and increases ethanol oxidation by a 70-80 fold.^35^ Individuals that carry the rs1229984 (A) allele, therefore, present a condition known as “alcohol flush reaction,” characterized by a visible flush on their face and body, and in some cases, symptoms such as nausea, headache, and general discomfort. Hence, carriers of the A allele tend to limit their alcohol intake. In line with our findings, a recent study by García-Martín *et al*.^36^ reported an inverse association between the reduced alcohol intake phenotype of the rs1229984 (A) allele and increased PD risk. Notably, although the effect of variant rs1229984 on alcohol metabolism is well understood, the mechanism by which alcohol intake reduces PD risk remains mostly unknown and warrants further investigation. Another strongly associated SNP with *drinks per week* was rs29001570 (see **Supplementary Materials**), located in the *Alcohol Dehydrogenase 5 locus* (*ADH5*), which has also been previously associated with alcohol intake.^37^ *ADH5* is known to be involved in formaldehyde metabolism but has low affinity for alcohol, so the mechanism by which it associates with alcohol consumption is not well understood.

We found that variants associated with being *a current smoker* in the *smoking cessation* GWAS were causally associated with reduced PD risk. Although the IVW result reached statistical significance OR = 0.39 (95% CI 0.22 to 0.69; p=0.001) per doubling of odds, the MR-Egger estimate suggested the presence of unbalanced horizontal pleiotropy, which means that there may be other (independent) pathways besides being a *current smoker* by which the genetic variants exert their effect on PD risk. Considering that very few SNPs were suitable proxies for being a *current smoker*, the confidence interval was too wide to rule out pleiotropic effects, despite estimates being consistent with those from IVW. Importantly, given that the discovery GWAS ignored time to cessation, the phenotype has a heterogeneous nature, making interpretation of these genetically derived estimates difficult. Larger sample sizes for both the *smoking cessation* and PD datasets will be needed in order to confirm our findings, as suggested by the lack of statistical power to detect moderate effect sizes for variants associated with *smoking cessation* (See **Table 1**). Importantly, the majority of smokers have attempted to quit smoking at least once in their lifetime^38^, and tobacco dependence is associated with longer smoking duration and failure of quitting attempts.^39^ Therefore, *being a current smoker* could be interpreted simplistically as individuals being unable to quit because they have a stronger genetic predisposition to smoking, whereas *former smokers* have a comparatively weaker predisposition and therefore cana quit successfully. Thus, our results support the hypothesis of smoking being a protective factor against PD.

**Table 1.** Number of variants, R^2^ and *F*-statistic and statistical power for each of the five phenotypes under investigation.

| Exposure | Total SNPs | $R^2$ | F-statistic |
| --- | --- | --- | --- |
| Age of smoking initiation | 7 | 0.00102 | 49.74375 |
| Smoking initiation | 96 | 0.00498 | 64.24512 |
| Drinks per week | 42 | 0.00603 | 136.02064 |
| Smoking cessation | 8 | 0.00105 | 72.13881 |
| Cigarettes per day | 29 | 0.01037 | 121.88111 |

Lastly, we observed a non-significant but consistent trend of a causal association between *smoking initiation* and PD. Previously, Grover *et al*.^40^ reported a strong protective effect of *ever being a regular smoker* against PD, using data from the largest GWAS on risky behaviors (Linnér *et al*.^41^) as the exposure dataset and the PD GWAS summary statistics available from the PDGene database (Lill *et al*.^42^) as the outcome. Based on our results, we conclude that even larger samples for both the exposure and PD datasets will be needed in order to validate these results.

Interpreting univariate MR results can be challenging because SNP instruments associated with the exposure of interest can exert pleiotropic effects on correlated risk factors. For instance, some alcohol SNPs are also associated with smoking behavior or BMI. However, discarding SNPs based on an observed association with correlated traits can greatly diminish statistical power to detect an association. By fitting pleiotropic risk factors alongside the exposure of interest in a multivariable MR framework, we were able to evaluate the direct effect that our exposure of interest has on PD risk. While the multivariable MR approach is conceptually useful to tackle these issues, in practice, it only provided meaningful results for the *drinks per week* trait, and it produced very wide confidence intervals for the smoking variables. Our results suggest that the protective relationship of alcohol intake (*drinks per week*) on PD remains consistent even after controlling for the (potential) pleiotropic effects of these SNPs on smoking behavior, obesity, and educational attainment. Thus, the causal association between alcohol intake and PD is unlikely to be the product of confounding via smoking behavior.

A handful of recent studies have attempted to examine the causal relationship between smoking and alcohol intake on PD risk using Mendelian randomization. Nalls *et al*.^27^ reported little evidence for a causal relationship between PD and current smoking status (OR=0.545; 95% CI=0.230-1.291; p-value=0.1681). Noyce *et al*.^28^ reported an inverse causal association between both current smoking status and increased alcohol intake and PD risk, but those associations were not explored in detail, and sensitivity analyses were not reported. Even though the Noyce *et al*. results and ours are similar, an important difference is the source of both the exposure (UK Biobank vs. Liu *et al*. 2019) and outcome (Chang *et al*. 2017 vs. Nalls *et al*. 2019) GWAS summary statistics datasets. The analyses presented here used datasets that comprised substantially larger and better-powered samples than those previously used. Also, we performed a series of sensitivity analyses to minimize the possibility of bias due to pleiotropy.

Limitations of the current study include the fact that we were not able to perform subgroup analyses, for instance, to determine whether the protective effect of alcohol intake on PD risk is the same in both men and women. That is an interesting question because, generally, men tend to drink more and more often than women. At the same time, we could not take into account the survival effects of alcohol intake and smoking behavior. It is known that both smoking and excessive alcohol drinking increase the risk of earlier mortality, and therefore might be under-represented amongst PD patients.^43,44^ Finally, given that the PD GWAS consisted of a case-control design, the effects of smoking and drinking behavior can only be interpreted in the context of risk of developing PD, but are not informative of the effect that these behaviors might exert on disease progression of individuals with a current PD diagnosis.

Finally, it is crucial to bear in mind that smoking and drinking pose serious health risks to individuals. Evaluation and translation of these epidemiological clues for therapeutics have been difficult and thus far unsuccessful. However, a better understanding of the underlying biology and their relation with PD risk could ultimately help delineate novel targets for prevention or treatment without the adverse health effects of smoking and drinking.

## METHODS

### Datasets

We obtained GWAS summary statistics from the GWAS & Sequencing Consortium of Alcohol and Nicotine use (GSCAN) consortium (Liu et al^26^ https://conservancy.umn.edu/handle/11299/201564), in which 556 genetic variants in 406 loci were associated with the following phenotypes: a*ge of smoking initiation, cigarettes per day, ever being a regular smoker, being a current vs. former smoker*, and the number of *drinks per week*, in sample sizes up to 1.2 million participants. The beta coefficients, standard errors, and p-values were extracted from all five datasets. In order to obtain a set of independent index SNPs for the MR analysis, each dataset was “clumped” using PLINK (http://zzz.bwh.harvard.edu/plink/clump.shtml). Variants selected as index SNPs were those with the smallest p-values <5e-8, while clumped SNPs were those in high linkage disequilibrium with the index SNP (R^2^ threshold of 0.001) or within a distance of 10,000 kb based on the 1000 Genomes project reference dataset. To evaluate whether the SNP instruments selected for each exposure were strongly associated with the exposure, the proportion of phenotypic variance for each trait explained by the SNPs (R^2^) and *F* statistics were calculated for the five traits (See **Table 1**). R^2^ for each trait was calculated based on the formula 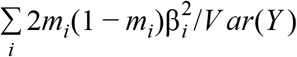 where *m*_*i*_ is the minor allele frequency and β_i_ the magnitude of the association between the i-th SNP on trait Y.^45^ The *F statistic* was calculated as previously described by Noyce et al^46^: 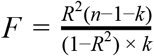, where *n* is the sample size and *k* the number of SNPs.

We also obtained summary statistics from the most recent PD GWAS from the International Parkinson’s Disease Genomics Consortium^27^ (IPDGC; http://pdgenetics.org/). This dataset consisted of 7.8 million analyzed genetic variants in 35,340 cases, 18618 UK Biobank proxy cases (first-degree relatives of PD patients), and 900,238 controls. The beta coefficients, standard errors, and p-values were extracted for those index SNPs previously identified in each study. For example, 7, 29, 96, 8, and 42 index SNPs were identified for the a*ge of smoking initiation, cigarettes per day, ever being a regular smoker, being a current vs. former smoker* (*smoking cessation*), and the number of *drinks per week*, respectively. We then inspected how many index SNPs were shared between PD and each of the five traits, in which all index SNPs for each trait were found on the PD dataset.

For the bidirectional-MR, 23 index SNPs were identified from the PD GWAS dataset and further extracted from the *cigarettes per day, ever smoking* and *drinks per week* GWAS datasets. Power calculations were undertaken using the method described by Brion et al.^47^ (https://shiny.cnsgenomics.com/mRnd/) in which the power to detect a true effect of OR 1.2 on PD risk in the iPDGC dataset with an alpha of 5% was evaluated for each trait (See **Table 1**).

### Effect allele harmonization

Harmonization across both the GSCAN and PD datasets was carried out as described by Hartwig *et al*., 2016^48^ prior to the MR analyses. First, all beta coefficients of the smoking phenotypes datasets were ensured to be positive (in order to facilitate interpretation of plots and as a requirement for further MR analysis). Negative beta coefficients were ‘flipped,’ e.g., they were multiplied by −1, the effect allele became the alternative allele, and the effect allele frequency was subtracted from 1. Then, we compared the effect allele coding in the smoking dataset and the Parkinson’s disease dataset and ensured that both had the same allele coding. Variants in the PD dataset that had different effect alleles in the smoking dataset were ‘flipped’ as previously described.

### Mendelian Randomization analyses

Before the MR analyses, we first harmonized both the GSCAN and PD datasets to align the effect alleles correctly. Four different MR methods were then selected to conduct the analysis: Inverse Variance Weighted (IVW)^49^, MR-Egger^50,51^, MR-PRESSO^52,53^, and GTCA-GSMR^25,53^, each with unique characteristics. A fundamental assumption required for MR inferences to be valid is that the instrument (SNP) and the outcome must be associated only through the exposure. If this assumption is violated, the presence of horizontal pleiotropy (an SNP associated with some other trait besides our exposure of interest) may bias MR estimates, potentially leading to erroneous conclusions.^20,22,54^ While the IVW model can only be interpreted under strong assumptions on pleiotropy^49^. The other three models exploit the statistical properties of MR estimates that are less vulnerable to these biases. For instance, MR-Egger relaxes this assumption and removes the constraint, in which the intercept corresponds to the average pleiotropy across SNPs and recalculates the causal estimate; if the intercept estimate differs from zero this is taken as an indication of unbalanced horizontal pleiotropy.^50,51^ MR-PRESSO identifies pleiotropic outliers under an IVW framework, and GTCA-GSMR calculates the causal estimate while identifying pleiotropic outliers. Applying these models in parallel can help compare MR estimates, making results more robust and less likely to be affected by weak violations of MR assumptions. We also generated funnel plots to inspect the presence of pleiotropic outliers manually, as asymmetry in the distribution of effect sizes indicate unbalanced horizontal pleiotropy. Cochran’s Q and I^2^ statistics were calculated to test for heterogeneity across estimates.^50,51^

### Multivariable MR

While our univariate MR analyses (i.e., fitting our exposure of interests one at a time) indicate whether genetic predisposition to these risk factors is associated with PD risk, it remains unclear whether other highly correlated environmental risk factors potentially mediate those relationships. To evaluate the direct effect between our exposures of interest and PD, we fitted a multivariable MR model by regressing out the genetic association between our SNP instruments (for each trait respectively) and the following putative risk factors in our MR analyses: BMI, years of education, smoking status and cigarettes per day (for the DPW MR only); alcohol intake (for the smoking-related trait MR only). For BMI (kg/m^2^) and education attainment (in SD units), we derived the genetic effect size estimates from the UK Biobank cohort. Data from the UK Biobank were QC-ed as per previous work^45^: 438,870 white British individuals were included in the GWAS analyses. The GWAS for BMI (UKB-FID: 21001) was conducted on 437,458 individuals and normalized educational attainment (UKB-FID: 6138) on 214,999 individuals using the BOLT-LMM^55^ mixed model software adjusting for recruitment age, sex and the first ten ancestral principal components. For alcohol and smoking-related traits, we adopted the effect size estimates from the GSCAN summary statistics itself. Genetic effect estimates of the SNP instruments (for each exposure) were then applied as covariates in the multivariable IVW MR model implemented via the MVMR R package.^56^

## Data Availability

We obtained GWAS summary statistics from the GWAS & Sequencing Consortium of Alcohol and Nicotine use (GSCAN) consortium (Liu et al https://conservancy.umn.edu/handle/11299/201564) and from the most recent PD GWAS from the International Parkinson’s Disease Genomics Consortium27 (IPDGC; http://pdgenetics.org/)

https://github.com/cdomingu/MRandPD

## ACKNOWLEDGEMENTS

We thank the research participants of 23andMe for their contribution to this study. CDB was supported by a Parkinson’s Foundation-APDA Summer Student Fellowship (PF-APDA-SFW-1913) and the UNAM “Programa 2019 para Actividades Especiales de Cooperación Interinstitucional (PAECI)”. XD’s work was supported by grants from the American Parkinson’s Disease Association. MER’s work was supported by the Australian National Health & Medical Research Council and the Australian Research Council through a NHMRC-ARC Dementia Research Development Fellowship (GNT1002821). CRS work was supported in part by NIH grants U01NS082157, U01NS095736, U01NS100603, and the American Parkinson Disease Association.

## AUTHOR CONTRIBUTIONS

MER and XD designed, conceived, and jointly supervised the study together with CRS. CDB and JSO performed and interpreted the statistical analyses. CDB drafted the first version of the manuscript with the support of MER, XD, and JSO. All authors assisted in the preparation and editing of the manuscript and approved its final version.

## DATA AVAILABILITY

The data that support the findings of this study are available from the corresponding author upon reasonable request.

## CODE AVAILABILITY

All code for this study is available at our GitHub repository: https://github.com/cdomingu/MRandPD.

